# Estimation of COVID-19 dynamics in the different states of the United States using Time-Series Clustering

**DOI:** 10.1101/2020.06.29.20142364

**Authors:** Fernando Rojas, Olga Valenzuela, Ignacio Rojas

**Affiliations:** Dpt. Computer Architecture and Computer Technology, CITIC-UGR, University of Granada, Spain; Dpt. Applied Mathematics, University of Granada, Spain

**Keywords:** COVID-19, Pandemic in the United States, Time Series, DTW distance, Hierarchical Clustering, SIR model

## Abstract

Estimation of COVID-19 dynamics and its evolution is a multidisciplinary effort, which requires the unification of heterogeneous disciplines (scientific, mathematics, epidemiological, biological/bio-chemical, virologists and health disciplines to mention the most relevant) to work together in a better understanding of this pandemic. Time series analysis is of great importance to determine both the similarity in the behavior of COVID-19 in certain countries/states and the establishment of models that can analyze and predict the transmission process of this infectious disease. In this contribution, an analysis of the different states of the United States will be carried out to measure the similarity of COVID-19 time series, using dynamic time warping distance (DTW) as a distance metric. A parametric methodology is proposed to jointly analyze infected and deceased persons. This metric allows to compare time series that have a different time length, making it very appropriate for studying the United States, since the virus did not spread simultaneously in all the states/provinces. After a measure of the similarity between the time series of the states of United States was determined, a hierarchical cluster was created, which makes it possible to analyze the behavioral relationships of the pandemic between different states and to discover interesting patterns and correlations in the underlying data of COVID-19 in the United States. With the proposed methodology, nine different clusters were obtained, showing a different behavior in the eastern zone and western zone of the United States. Finally, to make a prediction of the evolution of COVID-19 in the states, Logistic, Gompertz and SIR model was computed. With these mathematical model it is possible to have a more precise knowledge of the evolution and forecast of the pandemic.

## 1. Introduction

The COVID-19 epidemic started in Hubei Province, China, around December 2019. Since then, the disease has been spread to all continents and countries of the world, being categorized as pandemic by World Health Organization on March 11th.

In recent months, contributions have been made that analyze the evolution of different countries, implementing mathematical models to predict their evolution. Traditional predictive models for infectious diseases mainly include models for predicting differential equations and models for predicting time series based on statistics and random processes

For example, in [1] a methodology with the aim of estimating the actual number of people infected with COVID-19 in France is presented, since according to the authors, the number of screening tests carried out and the methodology do not directly calculate the actual number of cases and infection mortality rate (IFR). A mechanistic-statistical approach was developed that combines an epidemiological SIR model that describes this unobserved epidemiological dynamics, a probabilistic model that describes the data collection process and a method of statistical inference.

The logistic growth model, the generalized logistic growth model, the generalized growth model and the generalized Richards model were used to model the number of infected cases in the 29 provinces of China (and several countries), performing a a detailed analysis on the heterogeneous situations by four phases of the outbreak in China [2].

In [3] the Kermack-McKendrick SEIR model (Susceptible, Exposed, Infectious and Recovered) is presented to analyze the effects of behavioral changes on the reduction in community transmission in Mexico. A variable contact rate over time is proposed and the consequences of disease spread in an affected population of non-essential activities is analyzed.

The behavior of the virus in Japan has also been analyzed [4]. By February 29, 2020, in addition to the 619 confirmed cases (passengers and crew members) infected with COVID-19 in a cruise ship (near Tokyo), 215 locally transmitted cases had been also confirmed in Japan. To evaluate the effectiveness of reaction strategies based on avoiding large accumulations or crowded areas and to predict the spread of COVID-19 infections in Japan, in [4] a stochastic transmission model by expanding the epidemiological model based on SIR (Susceptible-Infected-Removed) had been presented. The simulation results showed that the number of Infected and Removed patients will increase rapidly if there is no reduction of the time spent in crowded zone.

In [5] using the Maximum-Hasting (MH) parameter estimation method and the SEIR model, the spread of COVID-19 and its prediction in South Africa, Egypt, Nigeria, Senegal, Kenya, and Algeria under three intervention scenarios (suppression, mitigation, mildness) is presented.

In addition to the most relevant epidemiological models used in the literature, models typically based on time series have also been used to analyze the behavior of the pandemic in different countries. The autoregressive integrated moving average (ARIMA) model is a mathematical model widely studied in the context of time series that is successfully applied in the field of health (estimate the incidence and prevalence of influenza mortality, malaria incidence, hepatitis, and other infectious diseases) as well as in different fields in the past due to its simple structure, fast applicability and ability to explain the data set. In [6] ten Brazilian states are analyzed using the autoregressive integrated moving average (ARIMA), the cubistic regression (CUBIST), the random forest (RF), ridge regression (RIDGE), the support vector regression (SVR) and the stacking-ensemble learning in the task of time series forecasting of the number of patient infected with COVID-19 with one, three, and six-days ahead. A forecasting model based on ARIMA has also been presented in [7] for Pakistan, presenting the high exponential growth in the number of confirmed cases, deaths and recoveries. In [8] ARIMA time series models were applied to forecast the total confirmed cases of COVID-19 for the next ten days using the model ARIMA (0,2,1), ARIMA (1,2,0) and ARIMA (0,2,1) for Italy, Spain, and France, respectively.

Currently, the analysis of the evolution of COVID-19 in America is of great importance due to the impact of this epidemic on this continent. In this contribution we will focus on the United States. The first patient detected in the United States was a travel-associated case from Washington state on January 19th, 2020. The preponderance of initial cased of infected patients with COVID-19 in the United States were correlated with travel to a ‘‘high-risk’’ country or close contacts of previously identified cases corresponding to the testing criteria adopted by the Centers for Disease Control and Prevention (CDC) (https://www.cdc.gov/). From March 1–31, 2020, the number of reported COVID-19 cases in the United States rapidly increased from 30 to 188,172, being the number of deaths from 1 to 5531, and detecting the virus all the states. At the end of April the number of infected reached 1069424 and the number of deceased stood at 62996. At the time of writing this contribution (14th june 2020) the number of infected is more than 2e+6 and more than 1e+5 deaths, being one of the countries of the world that is suffering with greater severity the disease of COVID-19.

In a recent paper [9] an attempt is made to estimate the actual number of infected people, even if they have not been counted. It was estimated that the true number of COVID-19 cases in the United States is likely in the tens of thousands, suggesting substantial undetected infections and spread within the country.

In [10] a relevant contribution is presented analyzing the sequence of nine viral genomes from early reported COVID-19 patients in Connecticut. From the phylogenetic analysis, it can be concluded that the majority of these genomes with sequenced viruses are from Washington State. By coupling their genomic data with domestic and international travel patterns, authors showed that early SARS-CoV-2 transmission in Connecticut was likely driven by domestic travel. The authors hypothesized that, with the growing number of COVID-19 cases in the United States and the large volume of domestic travel, new United States outbreaks are now more likely to result from interstate rather than international spread.

This contribution presents a methodology to analyze the evolution patterns of COVID-19 in the states of United States (including Puerto Rico and District of Columbia). A parametric similarity measure is presented, based on robust distance measure between time series, the dynamic time warping distance (DTW), with which the number of infected and dead in each of the states can be compared simultaneously, even though the start of the epidemic originated on different dates in each zone (therefore, the time series that need to be compared have different lengths).

To the best of our knowledge, this contribution is the first study that tries to develop a hierarchical clustering time series algorithm in order to globally compare and classify the behavior of all the states of United State simultaneously in their evolution of infected and deceased patients suffering COVID-19. Carrying out this classification is very useful, since it will allow to establish similarities and patterns in the evolution of the pandemic among the states of the United States. Once the different states have been grouped into a cluster (nine cluster are obtained), a SIR model, for each of the most representative elements of each cluster is analyzed. The simulation results of the nine SIR models evaluated are presented, indicating the most relevant parameters of the mathematical model and its prediction on the evolution of the pandemic in that state.

## 2. Material and Methods

A time series is a sequence of numerical (temporal) data points in successive order, which is naturally high dimensional and large in data size. There are two main operations that could be performed when working with time-series with its sequential data: a) the analysis of a single time series; b) the analysis of multiple time series simultaneously. This contribution is concentrated in the analysis of multiple time series for all the states of US suffering COVID-19, with the purpose of finding similarities between multiple time series by performing a clustering time-series methodology.

Clustering such complex objects is particularly advantageous because it may lead to the discovery of interesting patterns in time-series datasets, which contributes to a better understanding of the COVID-19 spread in different regions of the United States.

Clustering of time-series sequences has received noteworthy attention [11,12], not only as a formidable exploratory method and powerful tool for discovering patters, but also as a pre-processing step or subroutine for other tasks [13].

In this section, the database used is presented first (Section 2.1). Subsequently, a review of the most popular distance measures for time series is described (Section 2.2) and a new parametric distance is proposed. Then, existing approaches for clustering time-series data are briefly presented (Section 2.3) and the Logistic, Gompertz and SIR model, which will be used for modelling the time series of the most representative states in each of the cluster obtained, is described (Section 2.4).

### 2.1. Data set

The COVID-19 epidemic data set used in this contribution was collected from the Johns Hopkins University [14]. In this platform, the number of confirmed, deaths and recovered cases until June 21th 2020 for different countries are presented. For the United States, two additional .csv files are provided, in with detail of administration and province/state is reported (including Puerto Rico and District of Columbia). In order to compare countries behaviour, the time-series data are divided by state population.

### 2.2. Similarity/Distance measure in Time Series

In a simplified way, the similarity of two simple time series having the same number of points (denoted by m), and defined by X = {x_1_,x_2_,….x_m_} and Y = {y_1_,y_2_,….y_m_}, can be achieved by simply calculating the Minkowski (or Euclidean) distance (shortest path between two points) between points on both time series that happen at the same time. This distance is the measure of similarity, denoted as d(X,Y), and it is a function that takes both times series (X,Y) as input and calculates their distance “d”, defined as:

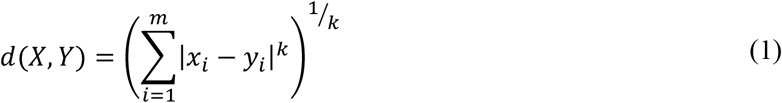

When k=2, the distance between two series is called Euclidean Distance. Using the Minkowski distance is a good metric to analyze the similarity of two time series, if these time series are synchronized (that is, all similar events in both time series occur at exactly the same time) and have the same length.

The evolution of time series in the different states of the United States present a different start date, both for the number of confirmed and death cases, and therefore its length is also different. Suppose by analogy the time series of the sound of a mother’s voice when she speaks slowly to her child. If the mother says the same phrase quickly, the child will most likely recognize that she is still his mother. However, if the Euclidean distance between both series were used as a metric, these two time series would have a very low similarity and would not be considered fundamentally equal. This would lead to the conclusion that the two voices did not come from the same person. To solve this problem, the dynamic time warping distance (DTW) method is frequently used in the bibliography [15].

DTW is a technique that can be considered as an extension of the Euclidean Distance between series [16], that calculates an optimal match between two given time series with certain restriction, performing non-linearly in the series (by stretching or shrinking along its time-axis). This distortion (denoted as warping) between two time series is used to find corresponding regions and determine the similarity between them.

The DTW of two series X and Y, defined as X = {x_1_,x_2_,….x_n_} and Y = {y_1_,y_2_,….y_m_} is computed in the following way. An *n-*by*-m* matrix D is computed with the (i.j)*th* element, defining the local distance of two elements by:

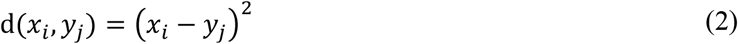

The point-to-point alignment between series X and Y can be represented by a time warping path W, defined as:

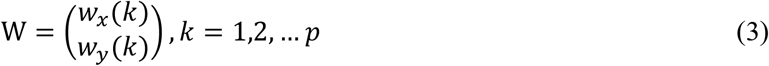

where *p* is the length of the warping path W, and w_x_ (k) and w_y_(k) represent the indexes in time series X and Y respectively. The warping path 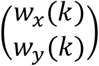 indicate that the w *th* element in time series X is *w*_*y*_(*k*) ^x^ mapping to the w_y_(k) *th* element in time series Y. There are some constraints and rules for the construction of the warping path:

- Every index from the first time series must be matched with one or more indices from the other time series (and vice versa)
- The first (the same for the last index) index from the first time series must be matched (not only this match) with the first (last) index from the other time series. That is, the warping path should start at W(1) = (1, 1) and end up at W(p) = (n,m).
- The mapping of the indices from the first time series to indices from the other time serie must be monotonically increasing, and vice versa. The adjacent elements of path W, W(k) and W(k + 1) must be subject to w_x_(k + 1) − w_x_(k) ≥ 0 and w_y_(k + 1) − w_y_(k) ≥ 0.
- The warping path should be also have the property of continuity, mathematically expressed as adjacent elements of path W, W(k) and W(k + 1) must be subject to w_x_(k + 1) − w_x_(k) ≤1 and w_y_(k + 1) − w_y_(k) ≤1.

The optimal match is denoted by the match that satisfies all the restrictions and the rules and that has the minimal cost, where the cost is computed as the sum of absolute differences, for each matched pair of indices, between their values. The DTW (minimal distance and optimal warping path) could be found using a dynamic programming algorithm:

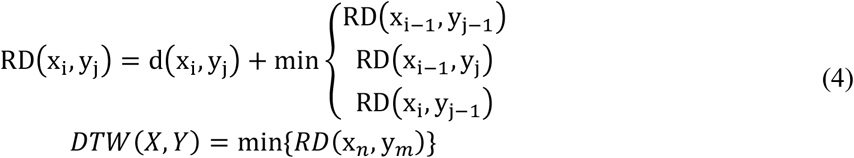

where RDx_i_, y_j_ is the minimal cumulative distance from (0, 0) to (i, j) in matrix D. In the methodology proposed in this paper, for each of the states analysed, both the time series of the number of infected and the time series of deaths will be simultaneously taken into account.

If each of these time series needs to be weighted differently, the following parametric metric, *DTW*_*∝*_(S_*A*_, S_*B*_) is defined:

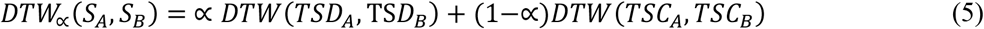

that measures the similarity in the evolution of the COVID time series for two states of the United States (S_A_ y S_B_), TSC_A_ and TSC_B_ represent the time series of the number of infected, TSD_A_ and TSD_B_ represent the time series of the number of deaths for the states S_A_ and S_B_ respectively. The parameter α (with 0≤α ≤1) indicates the relative relevance given to the similarity measure, taking into account the time series of infected or deaths.

### 2.3 Clustering method for time series

Clustering is a data mining technique in which similar data are divided into related or homogeneous groups, in an unsupervised way, that is, without knowing a priori advanced knowledge of the data. For the problem presented in this contribution, working with time series of states of the United States suffering COVID-19, given a set of individual time series data, the objective is to group similar time series into the same cluster.

The problem of grouping time series data is formally defined as, given a dataset of *N* time series data *Q*={*X*_*1*_, *X*_*2*_*… X*_*N*_}, find in an unsupervised way, a partition of *Q* into *K* cluster, denoted as *C*={*C*_*1*_, *C*_*2*_, *… C*_*k*_}, taking into account that homogeneous or similar series are grouped together based on a certain similarity/distance measure. In this paper, the parametric metric *DTW*_*∝*_(S_*AA*_, S_*B*_) is used and there is not intersection between clusters, therefore:

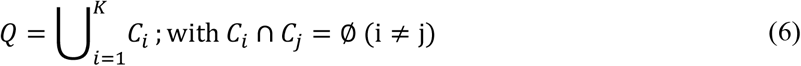

The methods used in the area of time series clustering [11, 17] are usually based in conventional clustering algorithm by substituting standard distance measurements with a more suitable distance to compare time series (raw methods) or converting series into normal data and using directly classical algorithms (Feature-based methods and models).

Among the most popular clustering algorithms, the hierarchical clustering and the k-means algorithm are widely used in time series clustering. In this contribution the hierarchical clustering is used, mainly due to its great visualization power and its simple and intuitive interpretation.

Hierarchical clustering creates a nested hierarchy of similar time series, according to a pair-wise distance matrix of the time series analyzed. The similarity measure *DTW*_*∝*_(S_*AA*_, S_*B*_) used is therefore essential in this time-series clustering process.

One of the most relevant characteristics of hierarchical clustering is its generality, since the user does not need to provide any parameters such as the number of clusters. As a disadvantage, hierarchical clustering has a high computational complexity when the number of elements to classify increases (the performance of hierarchical clustering is directly proportional to the squared size of the input data set). The methodology used to build the hierarchical clustering is the following:

1. - Calculate the distance between all the states of the United States using *DTW*_*∝*_(S_*AA*_, S_*B*_), for a certain value of the parameter α. This distance matrix, symmetrical and with the null diagonal, will be essential to analyze the similarity between the behavior of the different states.

2. - Search through the distance matrix in order to select the two most similar elements (in our case the time series of two states).

3.- Join (linkage) these two states to produce a new group that now have at least two objects (states).

4. - Update the distance matrix by calculating the distances between the new cluster and all other clusters.

5. - Repeat step 2 until all cases belong to a group.

The most widely used linkage criteria, such as single, average and complete linkage variants [18], were analyzed. Hierarchical clustering can be converted into a partitional clustering, with *k* cluster, by cutting the first *k* links.

### 2.3. Time series modeling

In this subsection, three models currently used in the bibliography for adjusting the evolution of COVID-19 from data will be briefly described.

#### 2.3.1. Logistic model

Mathematical models are formidable tools for understanding and predicting infectious diseases behaviour, being used in numerous viral diseases. A simple and easy-to-understand mathematical model is logistic regression analysis, used in modelling COVID-19 [19,20], expressed mathematically as:

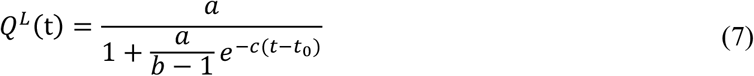

where *Q*^*L*^*(t)* is the cumulative cases of the logistic model at time *t* (can be the confirmed or dead patients), the parameters *a, b* and *c* are fitting coefficients of the model (numerical values of these three parameters depend on available data). Logistic models tend to under-estimate the total size of the infected/death population at the early stage, so they should provide lower bounds.

#### 2.3.2. Gompertz model

This model has been frequently used to describe the growth of animals and plants, as well as the number or volume of bacteria, virus and cancer cells [21]. In [22] this model was used to forecast the impact lethal duration of exposure on the mortality rates of COVID in seven countries (Germany, China, France, United Kingdom, Iran, Italy and Spain). It is based on sigmoid models fitted, that starts with an exponential growth and gradually decreases its specific growth rate, being a special case of the four parameter Richards model, and thus belongs to the Richards family of three-parameter sigmoidal growth models, using the following equation:

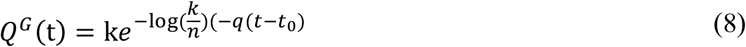

where *Q*^*G*^*(t)* is the cumulative cases of the Gompertz model at time *t* (can be the confirmed or deaths patients), n, k and q are fitting coefficients of the model, being q the parameter that modulates how the spreading rate is slowing down. These these three parameters depend on available data and must be adjusted using the available data from the COVID-19 time series, for each of the states of United States selected in the clustering process.

#### 2.3.3. SIR Model

Modelling the spread of infectious diseases usually performed by the categorization of the individuals in the population as belonging to one of several distinct compartments, which represent their health status with respect to the infection. In this paper the SIR model will be used, having three compartments: *S(t)* is the number of susceptible cases at time *t, I(t)* the number of infected cases and the function *R(t)* is the number of recovered persons in time *t* [23].

In order to understand and forecast the evolution of COVID-19 in the different states of the United States, the epidemic can then be analysed as the rates of transfer between these compartments [1], mathematically defined by the following non-linear systems of ordinary differential equations:

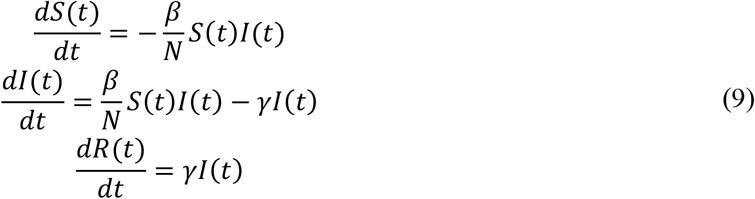

The parameter *β* and *γ* are the contact rate and average removal frequency respectively.

In order to solve the non-linear systems of ordinary differential equations presented in equation (9), initial conditions should be defined, being:

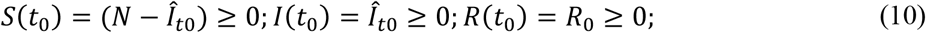

It follows from equation (9) that:

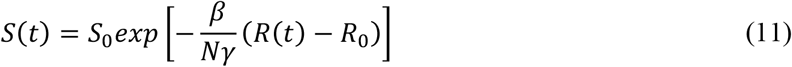

At the limit time *t* → ∞, assuming that the number of infected people is practically null, the number of susceptible people S(*t*_∞_) and recovered persons can be obtained as:

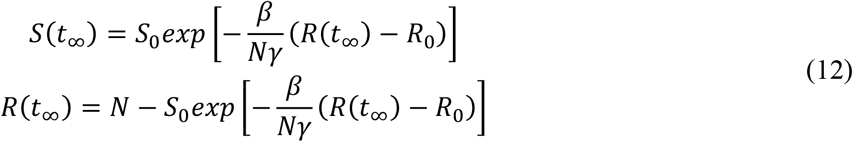

To model the behaviour of the epidemic using the equations presented in (7), the estimation of the parameters *β* and *γ* and the values of the initial conditions should be obtained from available data.

## 3. Results and discussion

To evaluate the performance of the proposed method, several experiments are conducted in this section for three values of the parameter α in the distance metric ***DTW***_*∝*_(***S***_***A***_, ***S***_***B***_). The time series of the states of the United States has been taken from John Hopkins database. For the computation of the distance metric, a threshold I_min_ has been defined, defining the minimum number of infected people to start the time series, being for this study I_min_=5 (the number of confirmed was greater than 5). Therefore, the length of the time series is different for each state, being on average 114 days (Figure 1). The index of each of the states is presented in Table 1.

**Table 1.** Distribution of the states obtained by means of hierarchical clustering with 9 clusters (α=0.5 and in bold, the state for which the SIR model is calculated)

| Cluster Number ($C_N$) | $D_{Cluster}$ | States ($\alpha=0.5$) |
| --- | --- | --- |
| 1 | 0.020 | (1) Arkansas, (2) **California**, (3) Florida, (4) Kansas, (5) Kentucky, (6) Maine, (7) Missouri, (8) Nevada, (9) North Carolina, (10) North Dakota, (11) Oklahoma, (12) South Carolina, (13) Tennessee, (14) Texas, (15) Utah, (16) Vermont, (17) Washington, (18) Wisconsin |
| 2 | 0.033 | (19) Alabama, (20) Arizona, (21) Colorado, (22) Georgia, (23) Indiana, (24) Iowa, (25) Minnesota, (26) Mississippi, (27) **Nebraska**, (28) New Hampshire, (29) New Mexico, (30) Ohio, (31) South Dakota, (32) Virginia |
| 3 | 0.017 | (33) Michigan, (34) **Pennsylvania** |
| 4 | 0.022 | (35) Delaware, (36) **Illinois**, (37) Louisiana, (38) Maryland |
| 5 | 0.022 | (39) Alaska, (40) Hawaii, (41) Idaho, (42) Montana, (43) **Oregon**, (44) Puerto Rico, (45) West Virginia, (46) Wyoming |
| 6 | 0 | (47) **District of Columbia** |
| 7 | 0 | (48) **New Jersey** |
| 8 | 0.035 | (49) Connecticut, (50) Massachusetts, (51) **New York** |
| 9 | 0 | (52) **Rhode Island** |

**Figure 1.**
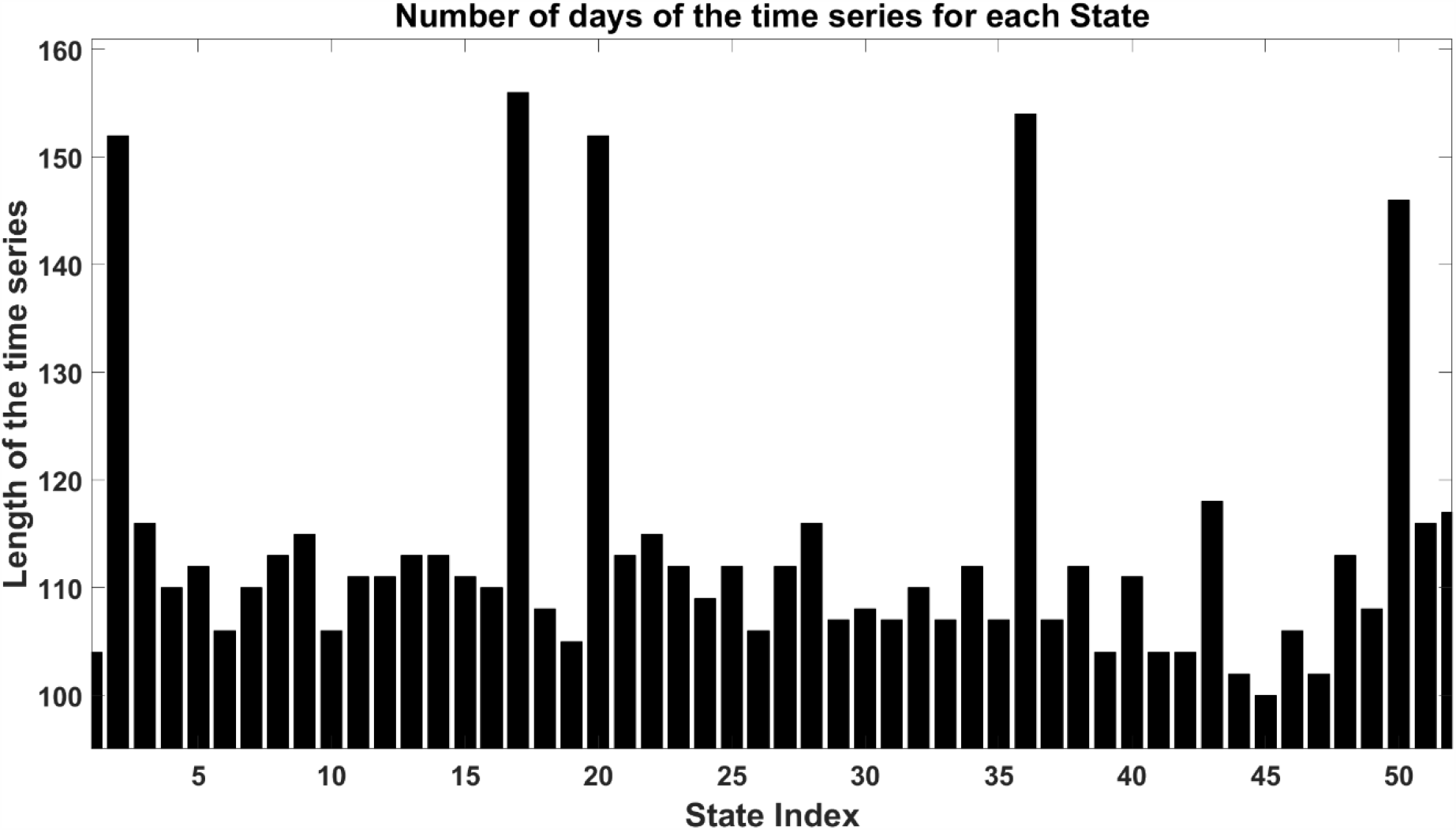
Length of the different time series, corresponding to the states analyzed.

The values of the parameter α analyzed will be {0,0.5,1}. This section of results begins with the value of α = 0.5, that is, the information of the confirmed patients time series having the same relevance as the time series of deaths for the final computation of the distance *DTW*_*∝*_(S_*AA*_, S_*B*_). The distance matrix between the different states is presented in Figure 2 (for a better visual representation, the distance matrix has been multiplied by a constant and the states are ordered according to the cluster to which each one belongs).

**Figure 2.**
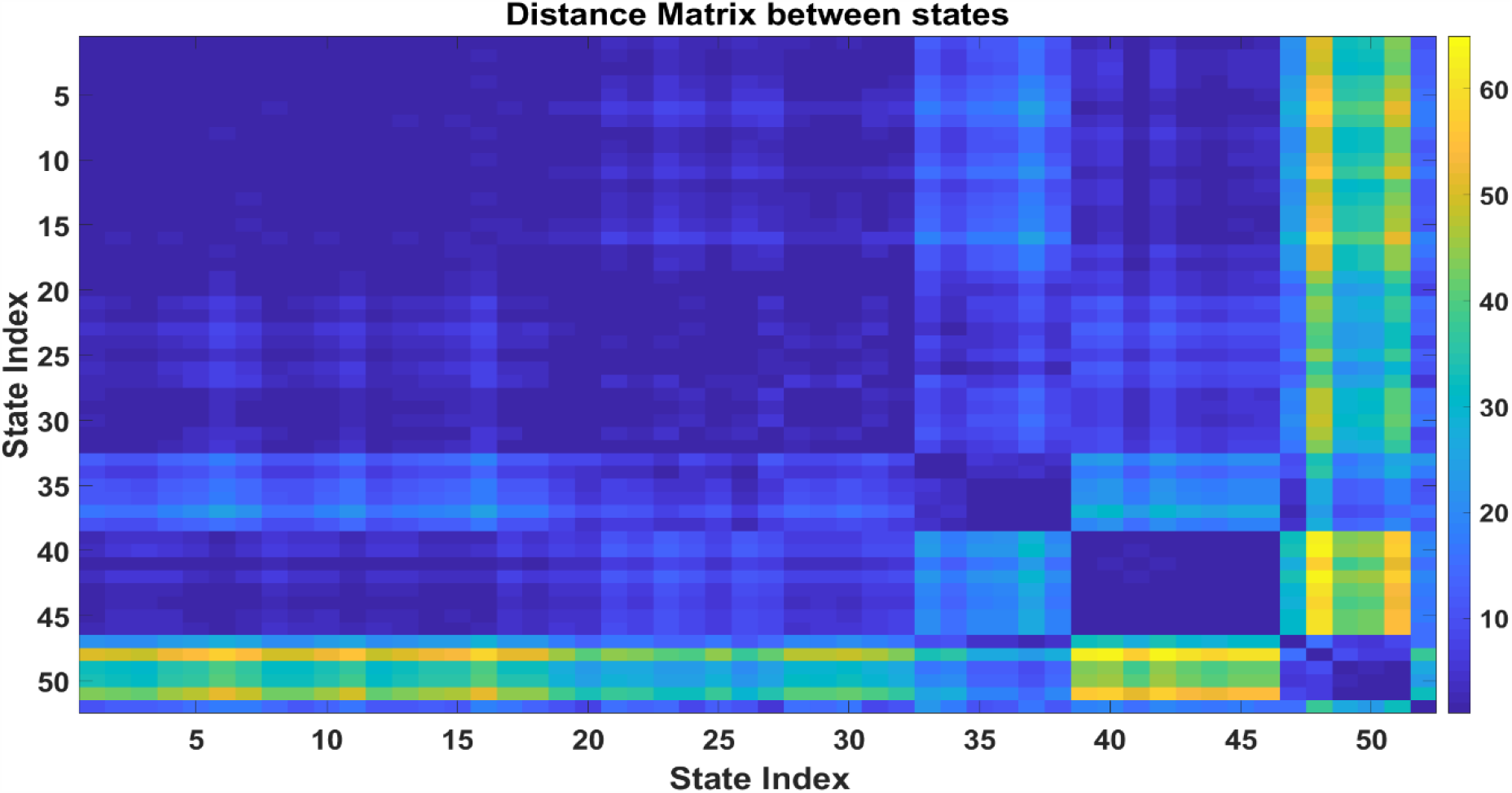
Distance or similarity symmetric matrix to characterize the behavior of the time series for the states of the United States (parameter α=0.5). The greater the similarity, the smaller the distance between the series (being the diagonal of this matrix of zero value).

The smaller the *DTW*_*∝*_(S_*A*_, S_*B*_) distance, the darker its representation. States that have a large distance (therefore have a low similarity in the behavior of their time series), are represented by yellow color (colorbar on the right side of the figure). To carry out the hierarchical cluster tree, average linkage is used. The average distance between all pair of states in any two clusters is defined as:

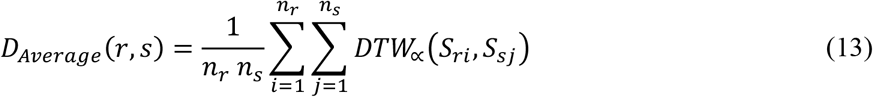

Where r and s represent clusters, and *n*_*r*_ and *n*_*s*_ is the number of states in cluster r and *s* respectively, being S_ri_ the *i*th state in cluster r and S_sj_ the *j*th state in cluster s. The hierarchical cluster tree obtained is presented in Figure 3.

**Figure 3.**
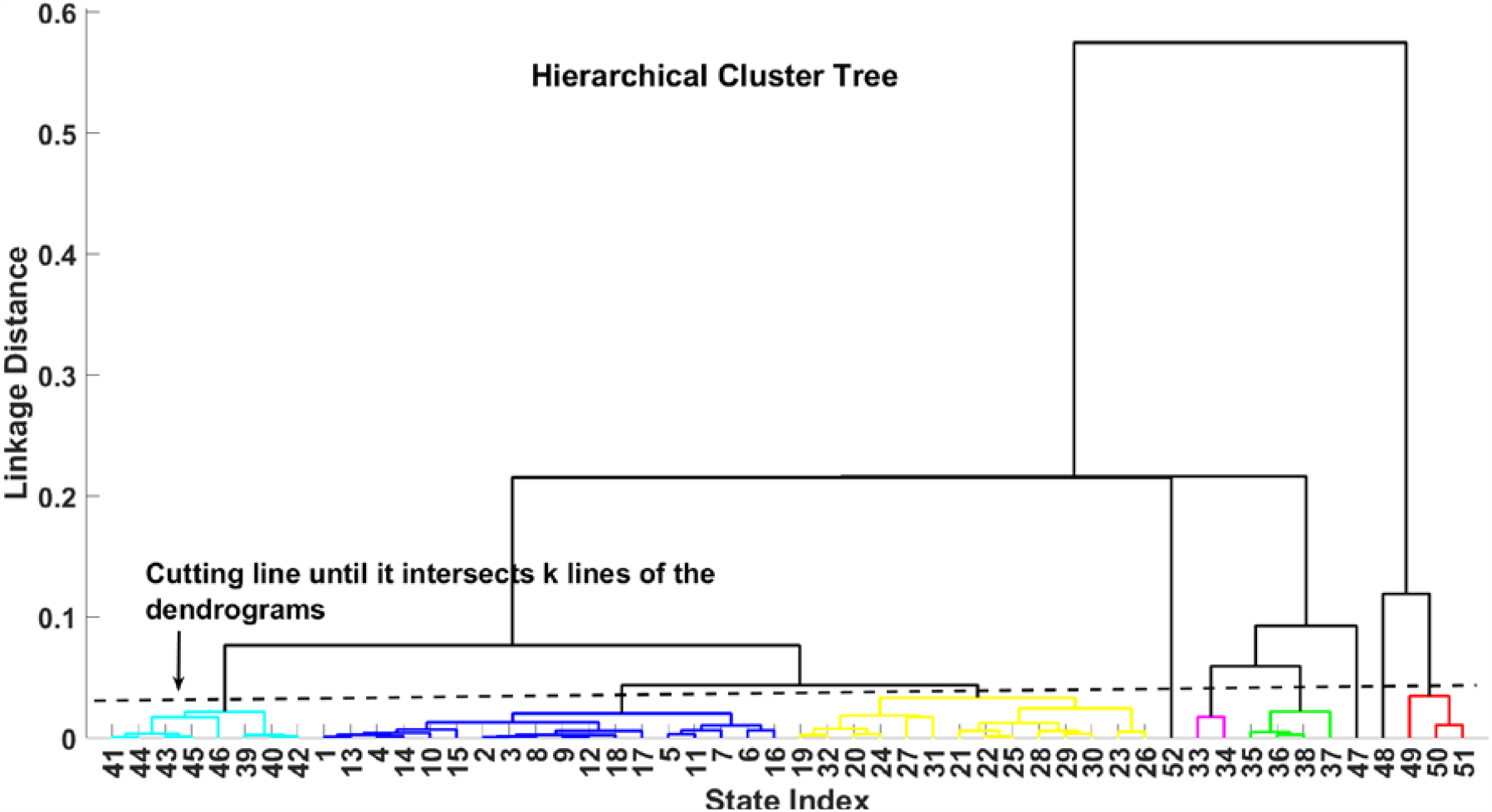
Hierarchical cluster tree obtained using as distance metric the *DTW*_*∝*_(S_*A*_, S_*B*_) and α=0.5

To analyse the accuracy of the obtained hierarchical cluster, the Cophenetic Correlation Coefficient (CP) is used [24].

The cophenetic correlation coefficient has been widely used in clustering problem, both as a measure of fitting degree of a classification to a set of data and as a criterion for evaluating the efficiency of various clustering techniques. For the problem presented in this contribution CP=0.90 using α=0.5.

The Calinski-Harabasz criterion (also denoted as the variance ratio criterion) is used to determine the optimal number, defined as:

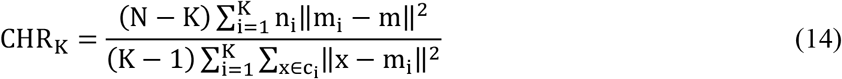

where the numerator quantifies the overall between-cluster variance, multiplied by (N-K), where N is the number of observations and K is the number of cluster. The denominator quantifies the overall within-cluster variance. The variable *m*_*i*_ is the centroid of cluster *i*, being *m* the overall mean of the sample data, *x* is a data point, *c*_*i*_ is the *i*th cluster and ‖m_i_ − m‖^2^is the Euclidean distance between two vectors. The larger the CHR_K_, the better the data partition (the clustering performed), therefore, the optimal number of clusters is obtained maximizing the Calinski-Harabasz criterion with respect to K. In the problem presented in this paper, the optimal number of clusters was 9 (using the parameter α=0.5) with the distribution shown in Table 1.

where D_Cluster_ is the distance between the elements that make up a cluster (its value is zero in the case that there is only one element in a cluster).

It is important to highlight the existence of various clusters with only one state (corresponding with District of Columbia, New Jersey and Rhode Island). Cluster 7 (New Jersey, listed as 48 in Table 1) links directly to cluster 8 ((49) Connecticut, (50) Massachusetts, (51) New York), which denote similar behaviour between these states. For cluster 6, (47)District of Columbia, the linkage is done for both cluster 3 ((33) Michigan, (35) Pennsylvania) and cluster 4 ((35) Delaware,(36) Illinois,(37) Louisiana,(38) Maryland). There are two large clusters (cluster 1 and cluster 2) that contain 18 and 14 states respectively, performing a direct linkage (meaning that these states have analogous performance). Its linkage is done through cluster 5, which contains eight states.

The similarities and distances between the different states and clusters obtained can be analysed using the results presented in the hierarchical clustering (Figure 3) and distance matrix (Figure 2).

Once the hierarchical clustering of the different states of the United States has been established and performed, it is relevant to model the time series (both infected and dead patients), using the models proposed in Section 2.3 (Logistic, Gompertz and SIR models). Tables 2 and 3 present the parameters of the Logistic and Gompertz models for all the states of the United States, for the time series of confirmed patients and deceased patients of COVID-19, respectively. The variable Adj_rsquare_ is degree-of-freedom adjusted coefficient of determination and the RMSE represents the Root Mean Squared Error (standard error, the difference between the actual data and the data obtained by the model)

**Table 2.**
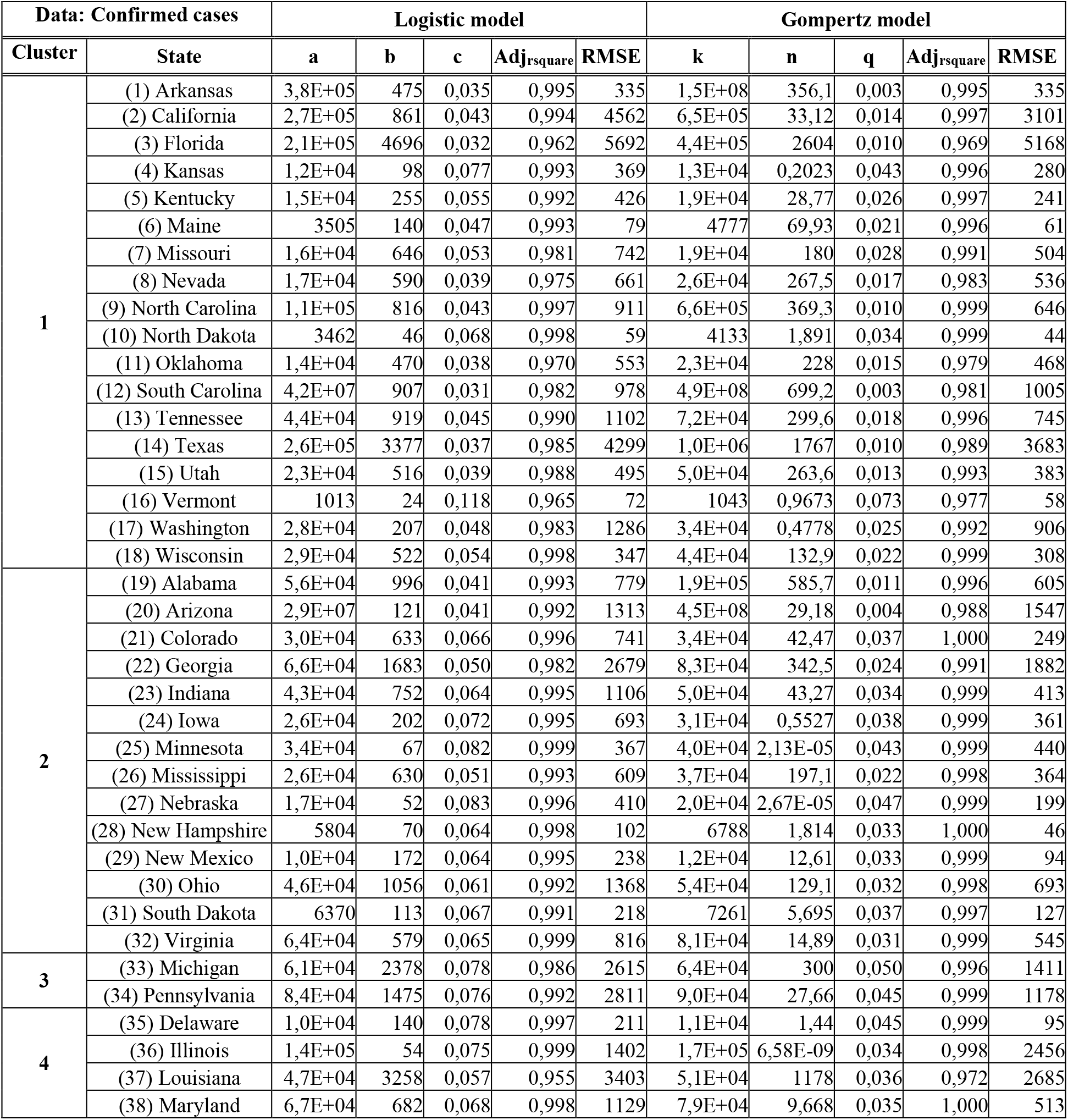
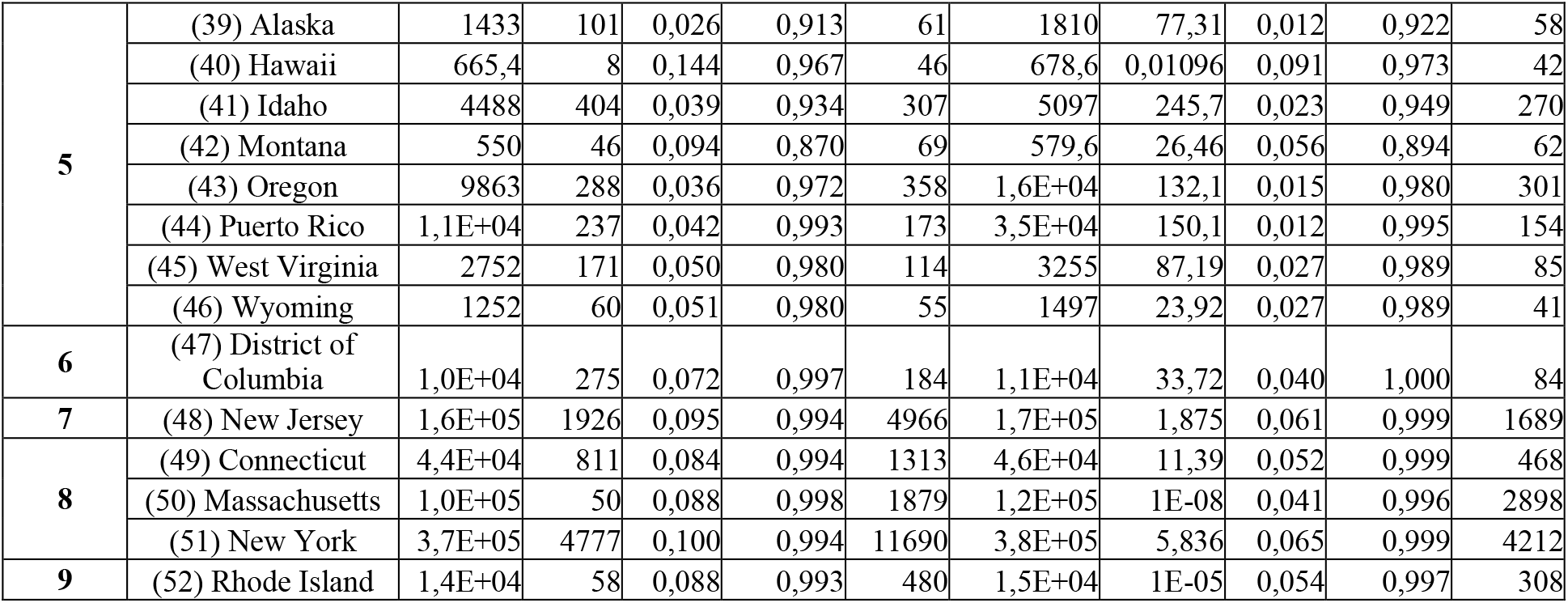
The prediction epidemic model results of COVID-19 for confirmed cases in all the states of United States, using Logistic and Gompertz model (the similarity metric α=0.5)

**Table 3.**
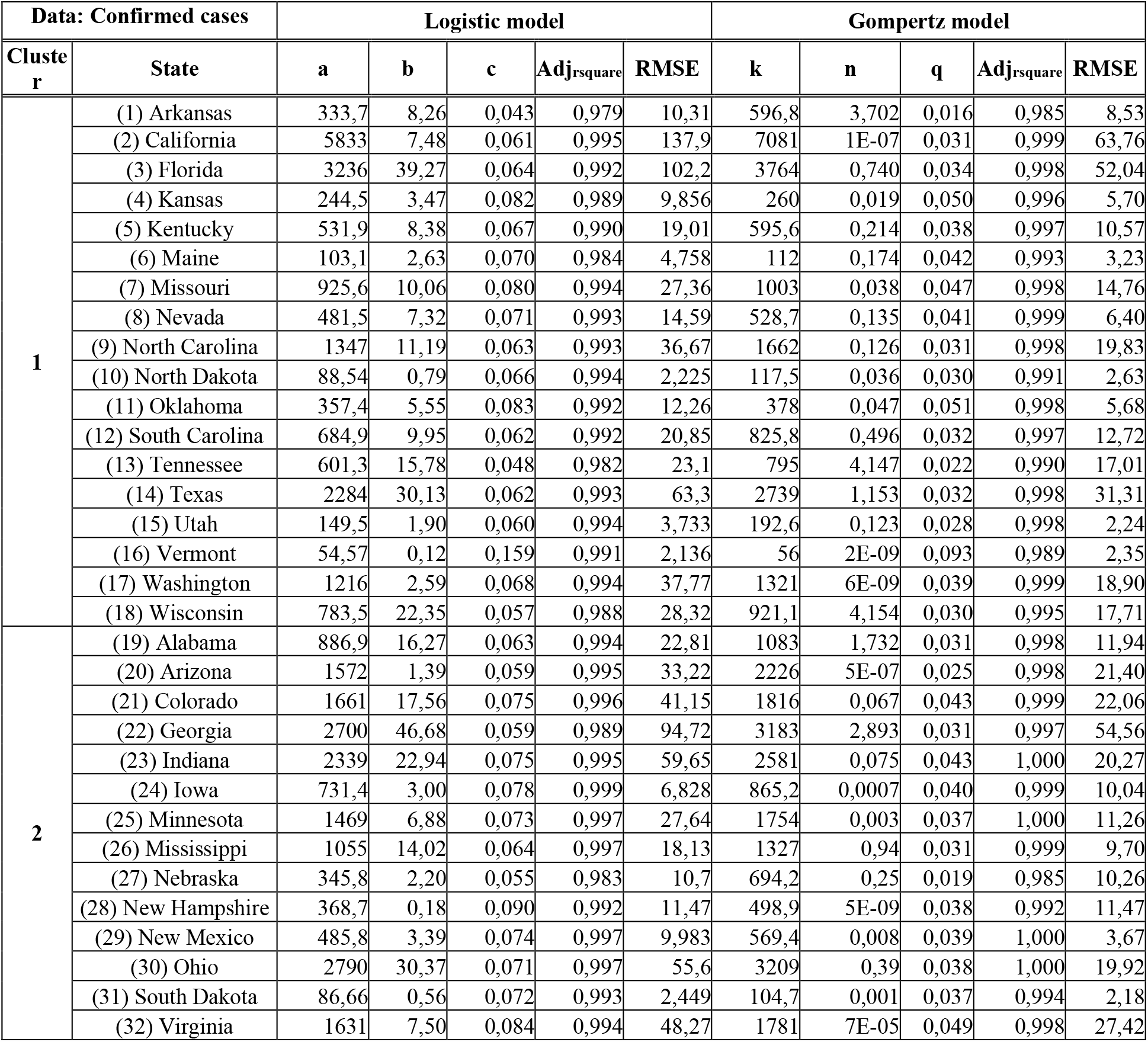
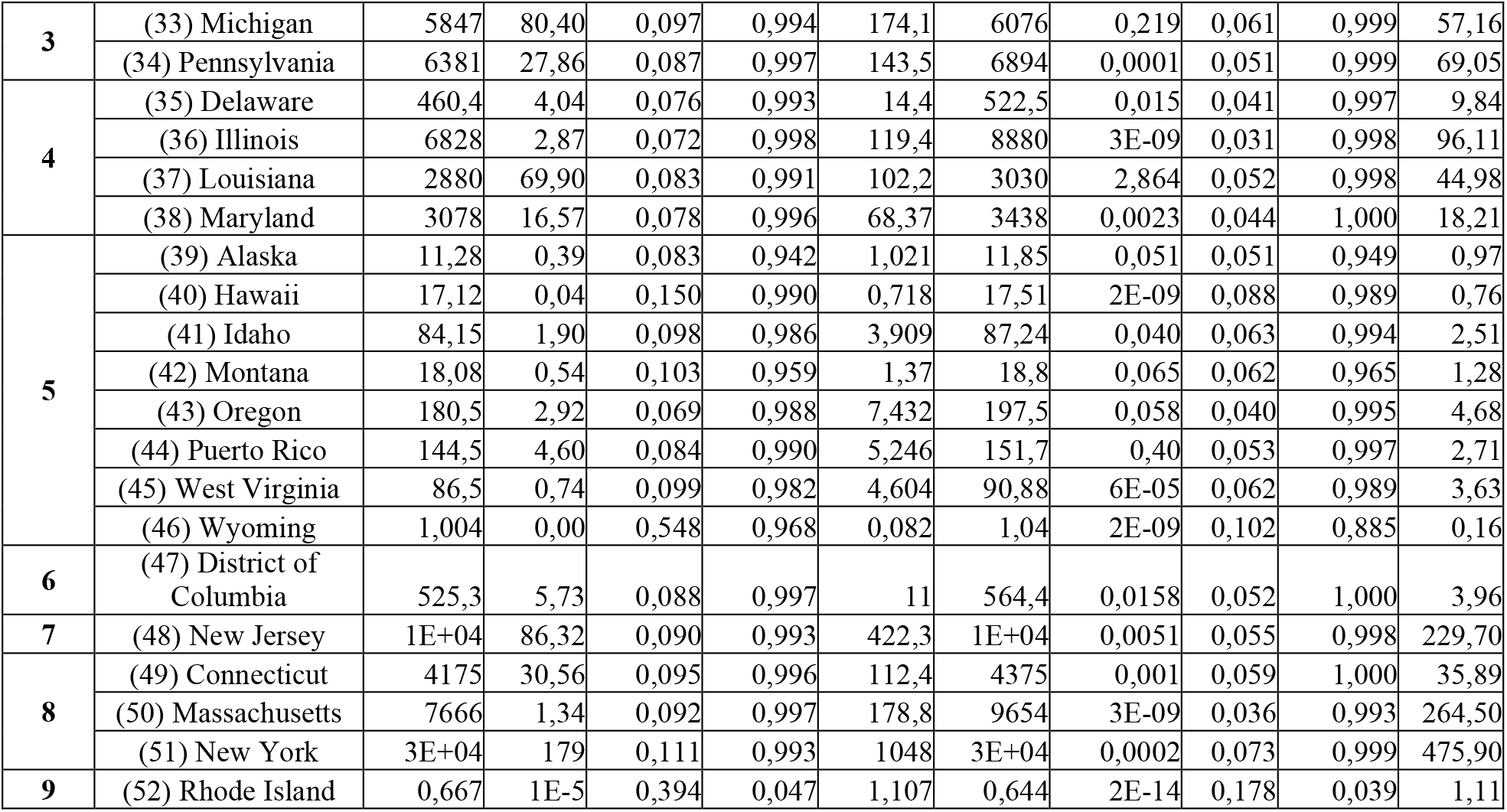
The prediction epidemic model results of COVID-19 for death cases in all the states of United States, using Logistic and Gompertz model (the similarity metric α=0.5)

Finally, the SIR model is calculated for the different representative states of each of the clusters obtained using the parameter α=0.5 (taking into account simultaneously both the number of infected and dead patients), being the parameters presented in Table 4. For the SIR model, the following variables are defined: C_N_ is the cluster number, N_D_ is the final number of days take into account for analysing the specific time series, *β* and *γ* are average contact and removal frequency used in equation (9). The parameter R is the reproduction number, i.e. number of people infected by a person with COVID-19, defined as:

**Table 4.** SIR parameters and predictions for representative state in each cluster.

| $C_N$ | State | $N_D$ | $\beta$ | $\gamma$ | $I_0$ | $R_0$ | $R$ | $T_{EE}$ | $N_{DE}$ |
| --- | --- | --- | --- | --- | --- | --- | --- | --- | --- |
| 1 | California | 148 | 1.615 | 1.576 | 36 | 1.025 | 0.995 | 26-nov-2020 | 342 |
| 2 | Nebraska | 110 | 0.173 | 0.083 | 25 | 2.075 | 0.487 | 08-Oct-2020 | 245 |
| 3 | Pennsylvania | 111 | 0.126 | 0.055 | 1653 | 2.297 | 0.399 | 17-Nov-2020 | 297 |
| 4 | Illinois | 148 | 0.146 | 0.065 | 33 | 2.228 | 0.447 | 12-Nov-2020 | 324 |
| 5 | Oregon | 117 | 0.172 | 0.15 | 110 | 1.144 | 1.036 | 05-Apr-2021 | 445 |
| 6 | District of Columbia | 101 | 0.17 | 0.106 | 192 | 1.605 | 0.619 | 07-Oct-2020 | 238 |
| 7 | New Jersey | 112 | 0.184 | 0.078 | 1011 | 2.354 | 0.322 | 10-Oct-2020 | 247 |
| 8 | New York | 116 | 0.187 | 0.075 | 2305 | 2.504 | 0.28 | 17-Oct-2020 | 257 |
| 9 | Rhode Island | 117 | 0.174 | 0.076 | 23 | 2.284 | 0.372 | 16-Sep-2020 | 230 |

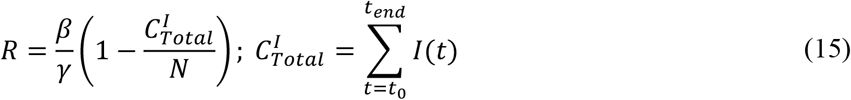

Being 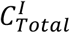 the total number of people infected at the end of the pandemic (t_end_) and N is the same constant presented in equation (9) (amount of susceptible population before the outbreak of COVID-19 in a specific state). R_0_ is the basic reproduction number, defined as the expected number of secondary cases produced by a single infection in a completely susceptible population and defined by:

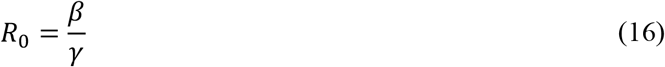

The variable T_EE_ is the time estimation of the SIR model in which the number of infected patients is very small, and therefore, it could be affirmed that the pandemic in that state would have ended. NDE denotes the number of days that the infection would have lasted in a given state. The prediction of the SIR model for the different state representative of the cluster obtained, analysing the evolution of total confirmed cases and novel cases per day, is presented in Figure 4.

**Figure 4.**
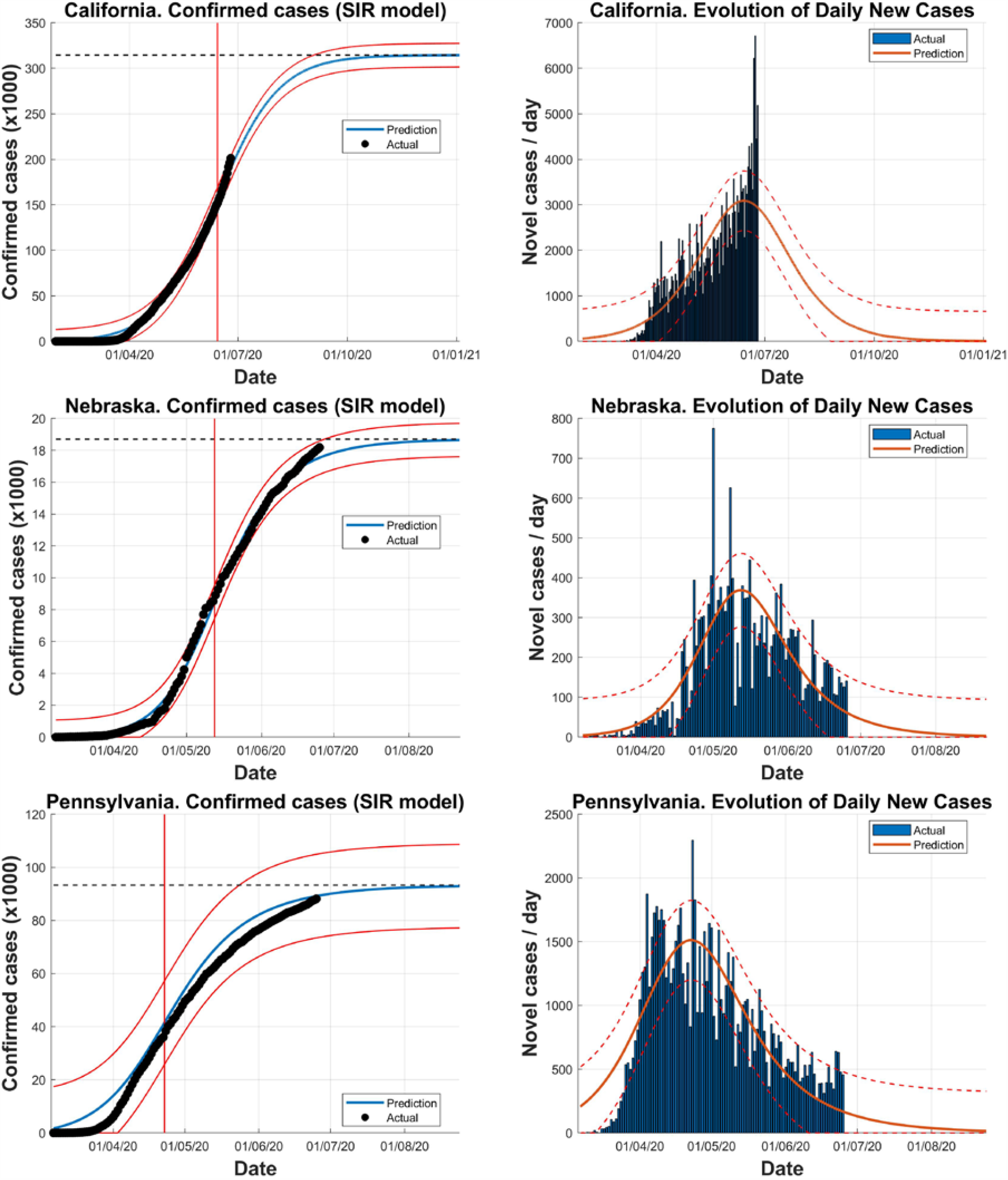
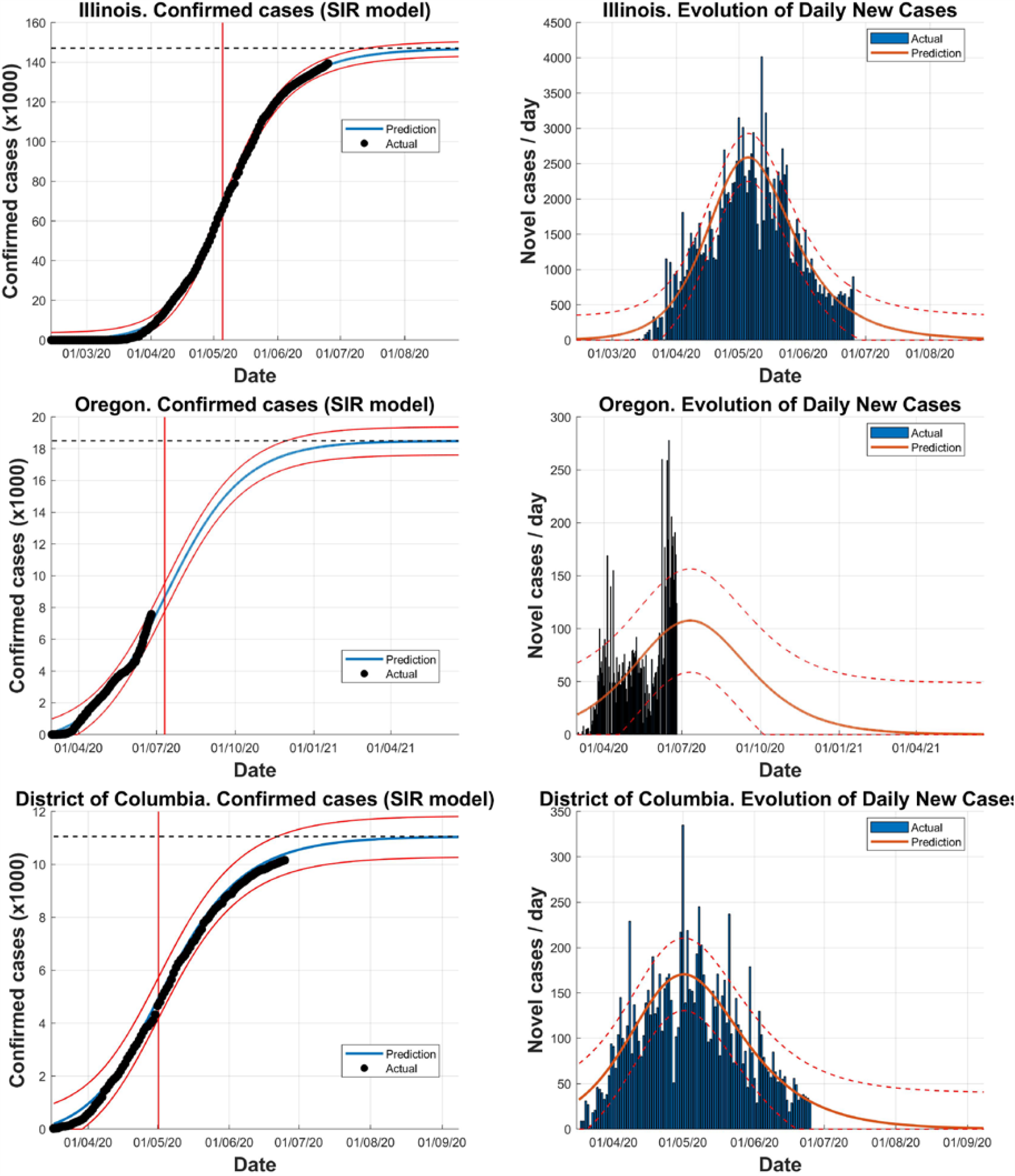
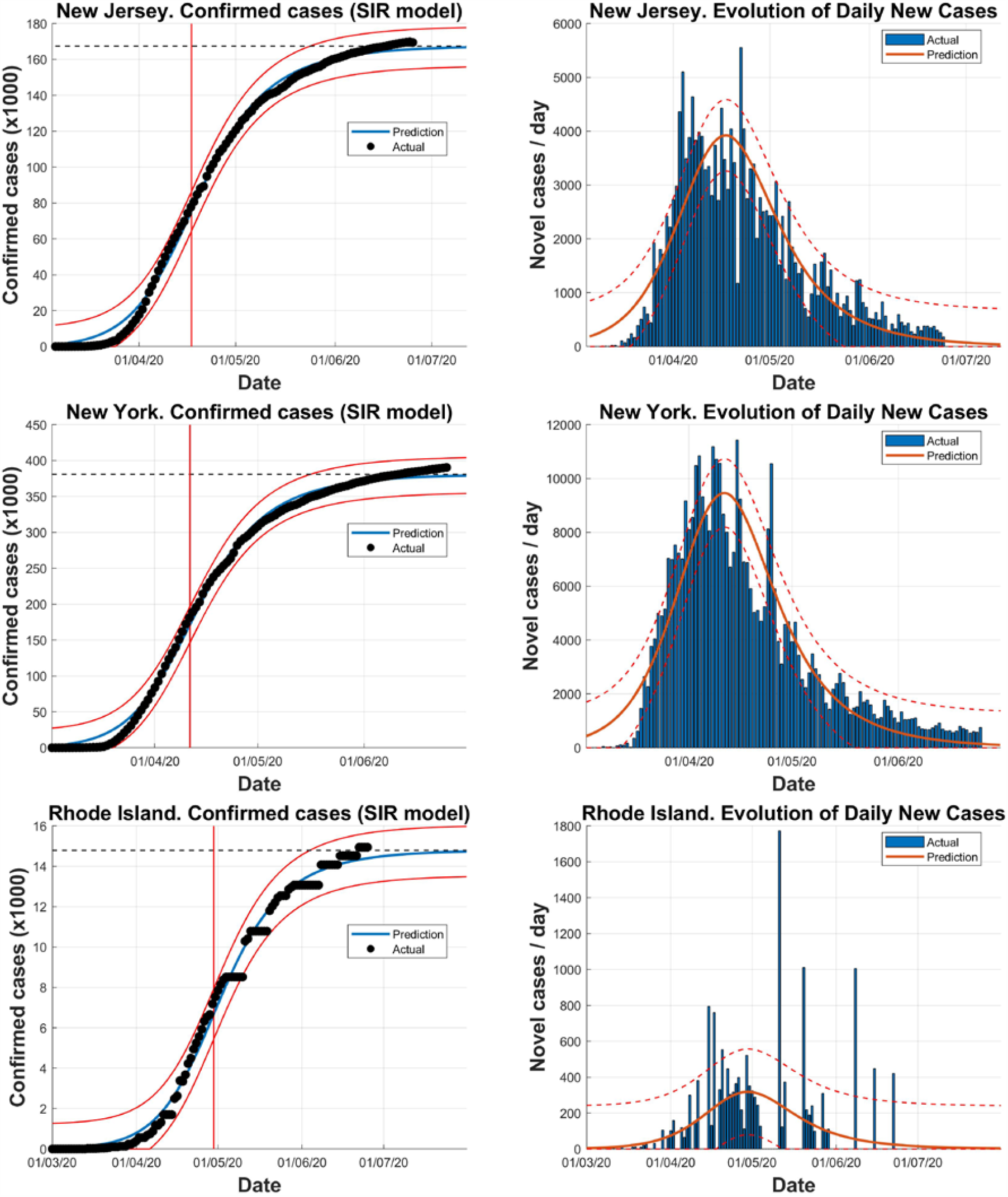
Prediction of the SIR model for the different states of the cluster obtained, analysing the evolution of total confirmed cases and novel infected cases per day.

## 4. Conclusions

A powerful tool for the analysis of time series is the grouping through clustering. Clustering time series is usually an unsupervised process, with the aim of finding behavioral similarities between the different time series that are analyzed. This article has proposed a parametric metric, based on the dynamic time warping distance, in order to measure the distance or similarity between time series corresponding to different states in the United States, taking simultaneously into account the behavior of the number of COVID-19 confirmed cases and deceased persons due to COVID-19. The proposed parametric metric, named *DTW*_*∝*_(S_*A*_, S_*B*_), is robust to the different lengths of data sequences (different beginning of the epidemic in the different states of the United States).

Using the Calinski-Harabasz criterion, the optimal number of clusters in which the different states of United States can be grouped was obtained, taken as value of α = 0.5 (same relevance for the time series of confirmed and death patients). A total of 9 heterogeneous clusters were found, in the sense that there are clusters within a large number of states (there are two large clusters, which encompass 18 and 14 states) and other clusters with only one state (indicating that their behavior has been unique, as they do not have excessive similarities with the rest of states).

Logistic, Gompertz and SIR mathematical models have been analyzed for the prediction and modeling of the evolution of the epidemic in the different states. For each of the clusters obtained, a representative state is selected and the SIR model was computed. This mathematical model, widely used in the bibliography, allows prediction of the evolution in a given state of the evolution of number of susceptible, infected and recovered patients, being this evolution and the estimate of the final size of the COVID-19 epidemic very relevant for the health authorities.

With the proposed hierarchical clustering procedure, it is possible to identify and summarize interesting patterns and correlations in the underlying data of the time series of the states of United States suffering COVID-19 and therefore determine similar behaviors that different states may have.

## Data Availability

data set used in this contribution was collected from the Johns Hopkins University

https://coronavirus.jhu.edu/map.html

## Acknowledgements

This contribution has been partially supported by the National Spanish project with reference RTI2018-101674-B-I00

